# Spatio-Temporal Distribution of HIV Cases in Ghana: A 5-Year Regional Assessment Using Routine Surveillance Data

**DOI:** 10.64898/2026.08.20.26360906

**Authors:** Osman Abdul-Fatawu Iddrisu, Frank Owusu-Sekyere, Abubakar Hudu Siddick, Bernard Kwadwo Yeboah Asiamah-Asare, Sylvester Dodzi Nyadamu

## Abstract

**Background:** The Human Immunodeficiency Virus and Acquired Immunodeficiency Syndrome (HIV/AIDS) remain a major public health concern in Ghana. Despite sustained progress in treatment and prevention, regional prevalence variations persist, driven by healthcare access, urbanization, and socio-economic factors. This study identifies trends and hotspots to guide effective HIV surveillance and control strategies in Ghana.

**Methods:** A retrospective ecological study was conducted using secondary HIV data confirmed by laboratory testing, from Ghana’s District Health Information Management System (DHIMS2) for the period 2020 to 2024. HIV prevalence was calculated as the number of confirmed cases per 100,000 population, using denominators from the Ghana Statistical Service 2021 Population and Housing Census. Spatiotemporal variation in prevalence was visualized using choropleth maps. Global Moran’s Index examined whether overall spatial dependency existed, followed by local indicators of spatial association (LISA), comprising local Moran’s I and the Getis-Ord Gi* statistic, to identify local clusters, outliers, and hotspots or coldspots.

**Results:** National HIV prevalence per 100,000 population rose from 0.68 in 2020 to 0.84 in 2024. The highest burden was in the southern and middle belt regions: Western North (2.16), Bono East (1.71), Eastern (1.30), Volta (1.06), and Ahafo (1.01). Northern regions remained consistently low throughout the study period, with Northern (0.32), Upper East (0.26), and Savannah (0.30) recording averages below 0.50 per 100,000. Global Moran’s Index indicated a dispersed pattern in 2020 (I = −0.43), spatially random pattern between 2021 and 2023, and weak positive spatial association in 2024 (I = 0.22).

**Conclusions:** Regional disparity in HIV prevalence in Ghana is widening, with greater burden concentrated in the more urbanized southern regions. Interventions guided by surveillance data and tailored to specific regions, including strengthened testing infrastructure and a more equitable distribution of health resources, are needed to curb transmission and support Ghana’s HIV and AIDS control programme.

## 1.0 Introduction

Human Immunodeficiency Virus and Acquired Immune Deficiency Syndrome (HIV/AIDS) remain among the most pressing concerns in global public health in the twenty-first century. The Joint United Nations Programme on HIV/AIDS (UNAIDS) and the World Health Organization (WHO) [1,13] estimated that approximately 39 million people were living with HIV worldwide in 2023, with about 1.3 million new infections and 630,000 deaths related to HIV recorded that year. Nearly two thirds of all people living with HIV, around 25.6 million, reside in Africa south of the Sahara [9,10]. Factors contributing to the higher prevalence in this region include limited access to health services, gender inequities, stigma, and inadequate provision of HIV prevention services [10,11,12].

The United Nations Sustainable Development Goals, under Target 3.3, commit to ending the AIDS epidemic and reducing new HIV infections and related deaths by 90% by 2030 [1]. Achieving this target will require sustained investment in prevention, treatment, and surveillance, particularly in countries with low and middle incomes, where health systems remain fragile. The economic consequences of HIV are considerable: each 10% increase in prevalence is associated with an estimated 2% reduction in national productivity [5,14,15], a relationship with substantial implications for countries where HIV prevalence continues to rise.

HIV remains a burden in Ghana, with consequences for both health and the economy. The disease affects not only individual health but also the productivity of the workforce and the demands placed on the health system [6]. According to the Ghana AIDS Commission, 334,095 people were living with HIV in Ghana in 2023, including 17,774 new cases and 12,480 deaths attributed to HIV [16]. The Ghana Health Service National HIV and AIDS Control Programme further reported more than 34,000 new infections between January and the third quarter of 2024 [3,4,17]. HIV is most common among individuals aged 24 to 39 years, with a growing number of infections recorded among university students and young adults [7]. Although the disease occurs across all regions, its distribution is uneven, and some regions consistently record higher case numbers than others. In 2016, adult HIV prevalence (ages 15 to 49) in Ghana was estimated at 1.6%, ranging from 2.7% in the Volta and Brong Ahafo regions to 0.7% in the Northern region [18]. These disparities reflect differences in population density, access to health care, socioeconomic conditions, and behavioural risk [2], and they underline why understanding how HIV cases are distributed across space and time matters for effective planning, early detection of outbreaks, and targeted intervention.

Existing studies have examined HIV epidemiology in Ghana [2–4], but most have been conducted at a single point in time [26], relied on behavioural proxies such as uptake of HIV testing [6,35], or offered a conceptual review without a rigorous spatiotemporal analysis of confirmed case data across multiple years [6,8]. Aboagye-Sarfo [20] modelled national trends in reported HIV incidence over time, yet few studies have examined spatial and temporal variation across all sixteen regions concurrently [6]. Regional disparity in HIV prevalence continues to challenge the achievement of national and global health targets, despite sustained efforts to curb transmission in Ghana [19]. This study addresses these gaps by applying spatiotemporal statistical methods to recent surveillance data from DHIMS2, using HIV prevalence per 100,000 population across all sixteen regions. The analysis examined the spatial distribution of HIV prevalence in Ghana between 2020 and 2024 and identified regions with persistently elevated or reduced burden, using measures of spatial autocorrelation and hotspot analysis.

## 2.0 Methods

### 2.1 Study Design and Data Sources

A retrospective ecological study was conducted using secondary data obtained from Ghana’s District Health Information Management System (DHIMS2) for the period 2020 to 2024. The ecological design examined HIV cases using population-level data aggregated by region and year rather than individual-level records. DHIMS2 is Ghana’s national routine health information system through which the Ghana Health Service (GHS) collects and reports routinely generated health information from health facilities across the country. For this study, DHIMS2 data were available for all 16 administrative regions of Ghana and comprised annual regional counts of HIV cases and laboratory-confirmed cases for 2020-2024. The data therefore represented aggregate routine health information rather than individual patient records. Population estimates for each region and year were used as denominators to calculate HIV prevalence and were obtained from the Ghana Statistical Service (GSS).

### 2.2 Study Area

The study was conducted in Ghana, West Africa, a lower middle income country with an estimated population of approximately 35 million in 2025, organized into sixteen administrative regions [21,22]. The population is predominantly young, with a median age of approximately 21 years. In HIV epidemiological research, individuals aged 15 to 49 years are conventionally defined as the sexually active population of reproductive age and therefore represent the principal population of interest [37]. See Figure 1.

**Figure 1.**
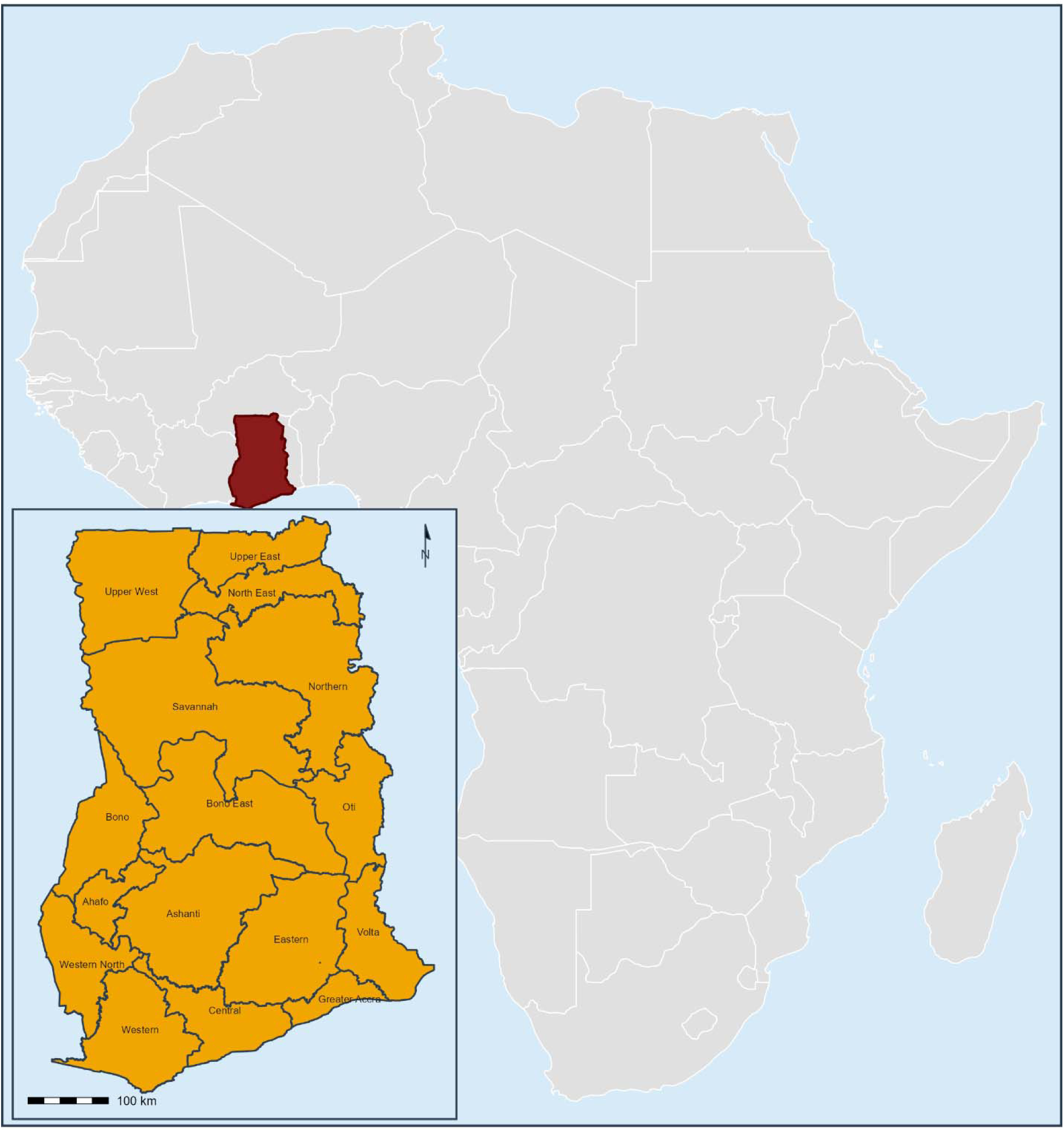
Map of Ghana showing all sixteen administrative regions

### 2.3 Study Population and Case Selection

The study population comprised all HIV cases confirmed through laboratory testing and reported to DHIMS2 by health facilities across Ghana’s sixteen regions between January 2020 and December 2024, irrespective of the age, sex, education, or socioeconomic status of the individuals affected. DHIMS2 records routine surveillance data as monthly counts at the facility level rather than as individual line-listed records; the analysis instead included the full set of monthly regional counts reported through the system for the study period, amounting to 960 region-month observations (sixteen regions across five years, twelve months per year), which were then aggregated into sixteen annual regional prevalence estimates for each year for the descriptive and spatial analyses that follow.

Cases were defined as HIV infections confirmed through laboratory testing under Ghana’s national HIV testing algorithm, which requires a reactive screening result to be confirmed by a second, distinct rapid test or, where indicated, a supplementary assay, before a diagnosis is recorded. DHIMS2 captures these diagnoses as standardized indicator counts submitted by facilities rather than as free text or diagnostic codes drawn from the International Classification of Diseases, and HIV status reported by patients themselves is not recorded as a confirmed case within the system. All confirmed cases (n = 54,467) meeting this definition and reported within the study period were included in the analysis. Records were excluded (n = 0) if they could not be assigned to one of the sixteen regions, if they represented duplicate facility submissions for the same month (n = 0) or if they fell outside the January 2020 to December 2024 reporting window (n = 0).

The period 2020 to 2024 was selected for three reasons. First, it represents the most recent five-year window for which DHIMS2 data are available in a consistent reporting format across all sixteen regions; changes to the reporting platform and to indicator definitions before 2020 make earlier regional data difficult to compare directly with the 2020 to 2024 records without extensive harmonization, which fell outside the scope of this study. Second, the period spans the COVID-19 pandemic and its immediate aftermath, an interval during which HIV service delivery, health-seeking behaviour, and case detection were widely disrupted; a temporally resolved spatial analysis of these years is therefore particularly relevant to understanding how the geographic distribution of the epidemic shifted under these conditions. Third, five years represents the minimum span needed to distinguish genuine trends in spatial clustering from ordinary year-to-year variation in regional counts, since both Global Moran’s I and the LISA statistics are sensitive to fluctuation within any single year. A longer time series extending back to 2015 would strengthen comparison between the period before the pandemic and the pandemic period itself, and we recommend this as a priority for future research.

### 2.4 Outcome measure

Monthly counts of confirmed HIV cases for all sixteen regions were extracted from DHIMS2. Regional HIV prevalence was estimated as the number of confirmed cases per 100,000 population. Prevalence, rather than incidence, was used as the primary indicator because DHIMS2 does not disaggregate newly diagnosed from previously diagnosed cases at the regional level in a format that permits valid computation of incidence across all sixteen regions. Prevalence per 100,000 population nonetheless captures meaningful differences in regional burden and tracked across five years, reveals the trends and spatial patterns central to this study’s objectives. Incidence-based analysis, ideally using facility-level line-listed data with dates of diagnosis, is recommended as a priority for future work.

### 2.6 Statistical Analysis

#### 2.6.1 Descriptive statistics and choropleth mapping of HIV prevalence

Descriptive statistics, including means and standard deviations, alongside the spatial and temporal analyses described below, were computed in R. Regional geographic boundaries were georeferenced and linked to the confirmed case data, then aligned with the sixteen administrative regions for each year from 2020 to 2024. Descriptive statistics were computed for regional HIV prevalence per 100,000 population for each year and for the study period. Choropleth maps were generated to show the spatial distribution of prevalence across Ghana’s sixteen regions for each year. Regions were classified using a quantile classification scheme with five class intervals, applied consistently across all five years to allow direct visual comparison of spatial patterns over time.

#### 2.6.2 Global spatial autocorrelation (Global Moran’s I)

Global spatial autocorrelation was assessed first, to establish whether overall spatial dependency existed in HIV prevalence across Ghana’s sixteen regions before any local-level cluster analysis was undertaken. Spatial relationships between regions were defined using a queen contiguity weights matrix, in which two regions were considered neighbours if they shared a common border or a common vertex [36]; this approach captures a broader and more realistic set of neighbouring relationships than rook contiguity and is particularly suited to the irregular shapes and varying sizes of Ghana’s administrative regions. The spatial weights matrix was standardized by row (style = “W”) prior to analysis [38]. Spatial correlation was quantified using the Global Moran’s I statistic [23], which measures overall spatial correlation across all sixteen regions simultaneously. Values of Moran’s I range from −1, indicating perfect dispersion, to +1, indicating perfect clustering, with values near zero indicating a spatially random pattern. Statistical inference for Global Moran’s I was based on a Monte Carlo permutation approach using 999 simulations, generating a reference distribution against which the observed statistic, its Z score, and pseudo p-value were evaluated [23].

#### 2.6.3 Local spatial clustering: local indicators of spatial association (LISA)

Local spatial patterns were examined using local indicators of spatial association, comprising local Moran’s I and the Getis-Ord Gi* statistic. The two measures were used together because they provide complementary information. Local Moran’s I identifies local clusters and spatial outliers, classifying each region into one of four categories, High-High, Low-Low, High-Low, and Low-High, with the latter two representing regions that differ markedly from their neighbours [39]. The Getis-Ord Gi* statistic served as a complementary, hotspot-focused measure, detecting statistically significant hotspots (clusters of high values) and coldspots (clusters of low values) without separately distinguishing outliers. Both statistics were computed using the same queen contiguity weights matrix, with statistical significance assessed through conditional permutation inference (999 simulations) at conventional confidence thresholds of 90%, 95%, and 99% [36,38]. All spatial analyses, Global Moran’s I, local Moran’s I, and Getis-Ord Gi*, were conducted separately for each year from 2020 to 2024 to track the temporal evolution of spatial dependency and clustering, using the R packages sf, spdep, ggplot2, and tmap.

## 3.0 Results

### 3.1 Descriptive Patterns of HIV Prevalence in Ghana, 2020–2024

HIV prevalence per 100,000 population rose in all sixteen regions between 2020 and 2024, from a range of 0.13 to 1.93 in 2020 to a range of 0.15 to 2.95 in 2024 (Figure 2). Nationally, average prevalence increased by 23.5% over the five years, from 0.68 to 0.84. The highest burden was concentrated in the southern and middle belt areas of the country. Prevalence rose in every region, most markedly in Western North, which increased from 1.80 in 2020 to 2.95 in 2024, a rise of 63.9%. Bono East ranged from 0.98 in 2020 to 1.50 in 2024, peaking at 2.14 in 2021. Eastern rose from 0.98 in 2020 to 1.48 in 2024, an increase of 51%. Rates remained consistently high in Greater Accra, Ashanti, Bono, and Ahafo, ranging from 0.44 to 1.37. By comparison, prevalence in the northern regions stayed consistently low: North East averaged 0.45 (range 0.10 to 1.20), while Savannah and Northern averaged 0.45 and 0.44 respectively, and Upper West and Upper East ranged from 0.17 to 0.50; Central and Oti recorded intermediate prevalence, at 0.65 to 0.92 and 0.17 to 0.50 respectively. These patterns point to a gradient running from north to south, with a disproportionate rise in prevalence concentrated in the more urbanized southern regions. Full summary statistics for each region across the five-year period are presented in Table 1.

**Figure 2.**
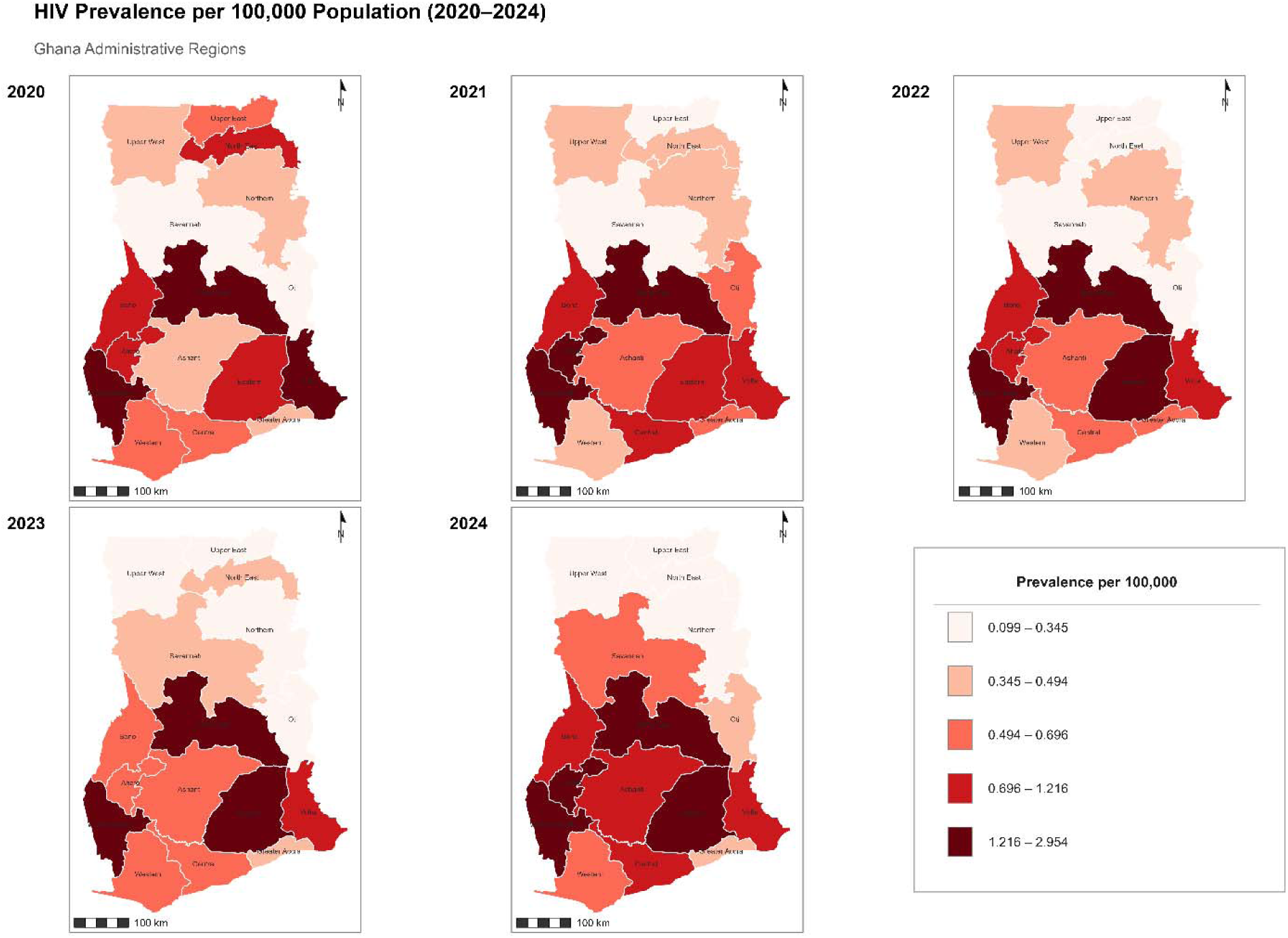
HIV prevalence maps (cases per 100,000 population) across Ghana’s sixteen administrative regions, 2020-2024

**Table 1.** Summary statistics of HIV prevalence per 100,000 population by region, 2020–2024.

| Region | Mean | Std. Dev. | Min | Max |
| --- | --- | --- | --- | --- |
| Western North | 2.16 | 0.56 | 1.49 | 2.95 |
| Bono East | 1.71 | 0.33 | 1.30 | 2.14 |
| Eastern | 1.30 | 0.26 | 0.98 | 1.60 |
| Ahafo | 1.01 | 0.34 | 0.62 | 1.37 |
| Volta | 1.06 | 0.19 | 0.78 | 1.28 |
| Bono | 0.81 | 0.23 | 0.51 | 1.14 |
| Central | 0.73 | 0.12 | 0.63 | 0.92 |
| Ashanti | 0.59 | 0.10 | 0.44 | 0.72 |
| Western | 0.52 | 0.12 | 0.35 | 0.66 |
| Greater Accra | 0.51 | 0.06 | 0.45 | 0.57 |
| North East | 0.48 | 0.42 | 0.10 | 1.20 |
| Upper West | 0.34 | 0.08 | 0.20 | 0.41 |
| Oti | 0.33 | 0.14 | 0.17 | 0.50 |
| Northern | 0.32 | 0.10 | 0.15 | 0.43 |
| Savannah | 0.30 | 0.16 | 0.13 | 0.50 |
| Upper East | 0.26 | 0.14 | 0.17 | 0.50 |

*Std. Dev.* = standard deviation; *Min* = minimum; *Max* = maximum

### 3.2 Global Spatial Autocorrelation of HIV Prevalence (Global Moran’s I)

In 2020, Global Moran’s I (Figure 3) indicated a dispersed spatial pattern (I = −0.427, Z = −1.77, pseudo p = 0.076), a result significant at the 90% confidence level and suggestive of a pattern unlikely to have occurred by chance. Moran’s I remained close to zero in each year from 2021 to 2023 (approximately −0.02, p > 0.81), indicating a largely random spatial distribution of HIV prevalence across regions during this period, with no clear evidence of clustering or dispersion. By 2024, Moran’s I had risen to 0.215 (Z = 1.55, pseudo p = 0.121), a shift toward weak positive spatial association that did not reach statistical significance at conventional thresholds but is consistent with an emerging clustering trend. Although the global evidence for 2024 was modest, the local Moran’s I and Getis-Ord Gi* analyses reported below identified local clustering patterns consistent with a spatial trend of increasing HIV prevalence over time.

**Figure 3.**
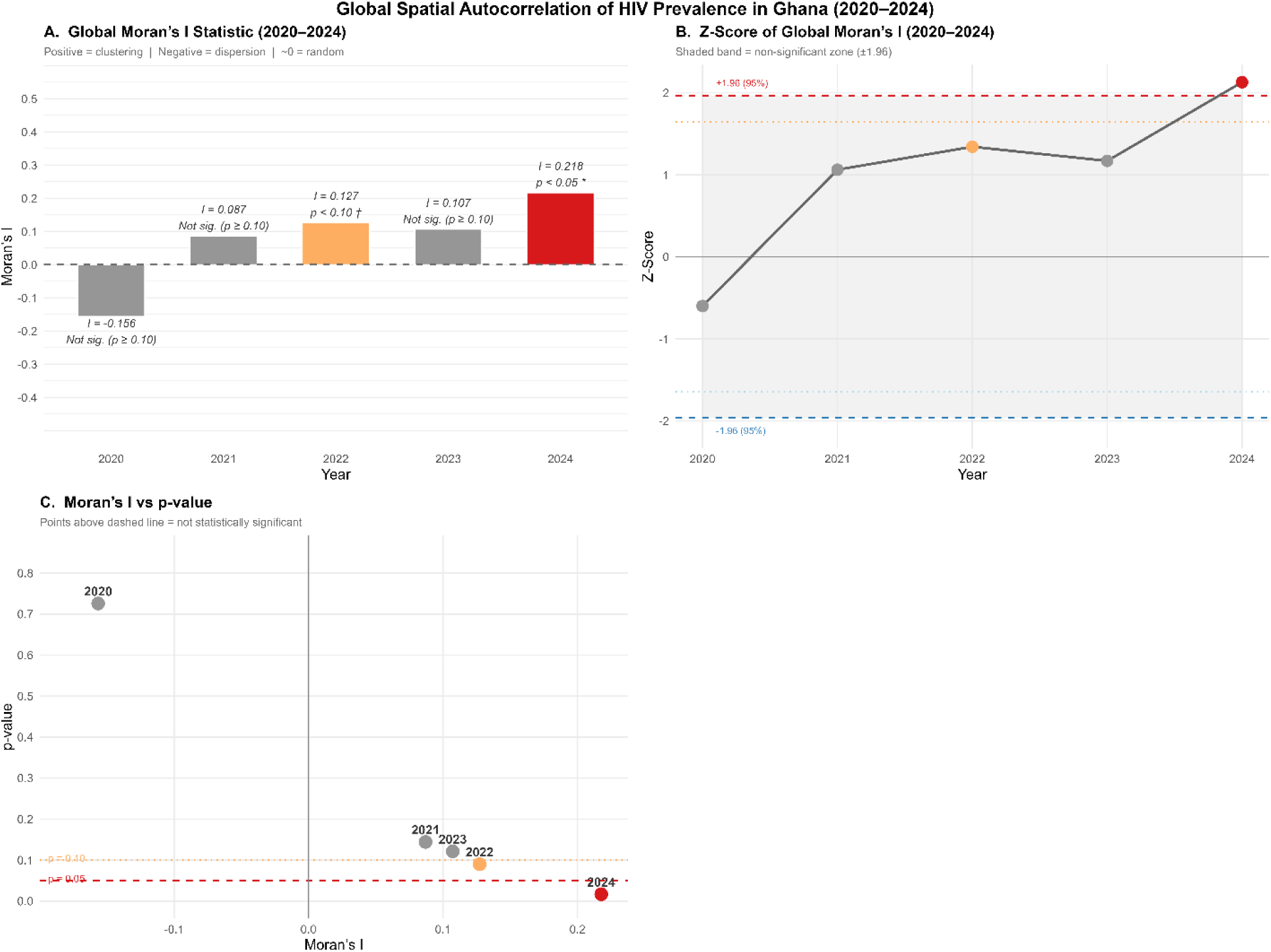
Global Moran’s I statistics for spatial autocorrelation of HIV prevalence in Ghana, 2020-2024

### 3.3 Local Hotspot Analysis (Getis-Ord Gi*), 2020–2024

The Getis-Ord Gi* hotspot analysis (Figure 4) revealed a distinct and continuing spatial trend in HIV prevalence intensity that reinforces the Global Moran’s I results reported above. No statistically significant hotspots or coldspots were detected in 2020. By 2021, a hotspot had emerged in Ahafo, while coldspots appeared across the northern belt, Northern, North East, and Upper East, together accounting for about 31% of the sixteen regions. In 2022, the coldspot extended further south to include Savannah. Hotspot intensity strengthened in 2023, with Ashanti emerging as a notable cluster at the 90% confidence level. By 2024, hotspots had shifted westward to Western North, Western, and Ahafo, at confidence levels ranging from 90% to 99%, while the northern coldspot persisted and intensified slightly, still covering about 31% of the country’s regions. Overall, the Getis-Ord Gi* results show that southern hotspots remained high-burden clusters in urbanized areas across four of the five study years; Ahafo, for example, was classified as a hotspot in 80% of the years examined.

**Figure 4.**
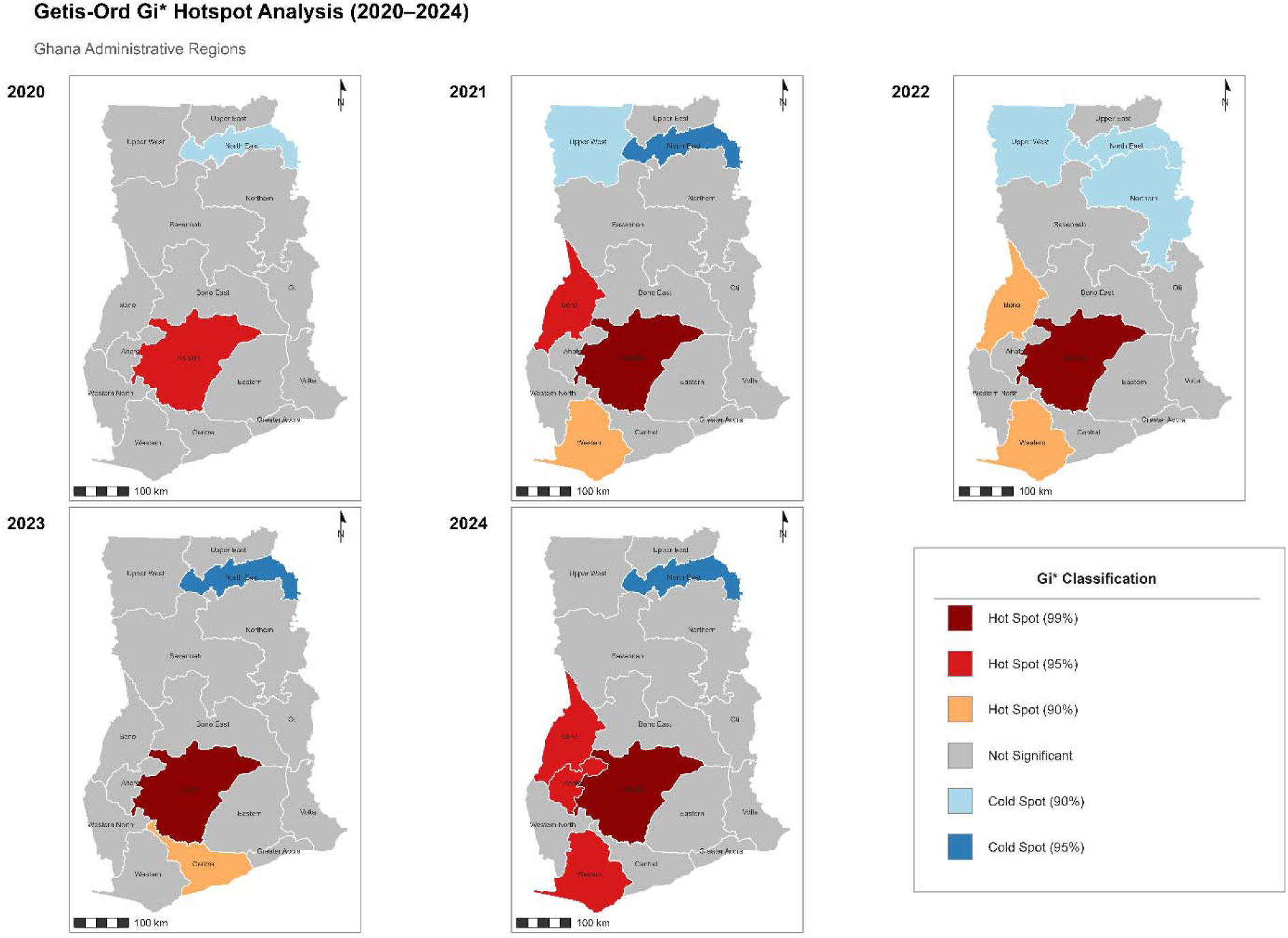
Getis-Ord Gi* hotspot and coldspot classification of HIV prevalence across Ghana’s regions, 2020-2024

### 3.4 Local Spatial Clustering and Outliers (Local Moran’s I), 2020–2024

Local Moran’s I analysis (Figure 5) identified persistent clustering patterns across the study period. High-High clusters were observed in Ashanti and Greater Accra, covering 12.5% of regions, and in Eastern, covering 6.0% of regions, throughout all five study years, suggesting a relatively stable and geographically concentrated transmission network.

**Figure 5.**
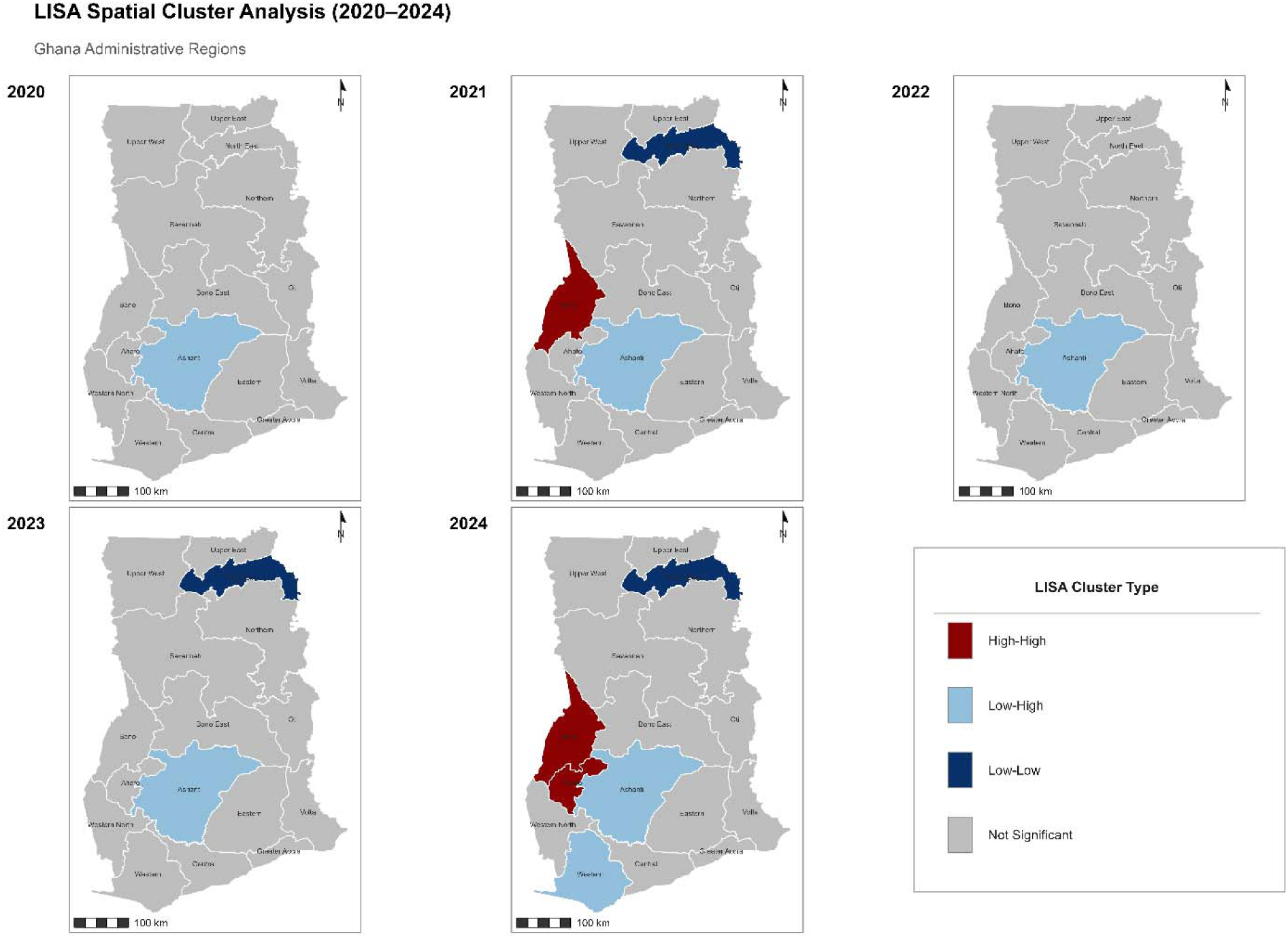
Local Moran’s I (LISA) cluster classification of HIV prevalence across Ghana’s regions, 2020-2024

Low-Low clusters, by contrast, dominated the northern belt, Northern, Upper East, Upper West, North East, and Savannah, persisting from 2020 through 2023 before spreading to additional areas in 2024. Western was classified as a High-Low outlier in four of the five years, meaning it recorded higher prevalence than its lower-prevalence neighbours, while Central and Volta were classified as Low-High outliers in most years, meaning they recorded lower prevalence than their higher-prevalence neighbours. These outlier regions may be particularly vulnerable to spillover transmission from hotspot regions to the south. These findings are summarized in Table 2.

**Table 2.**
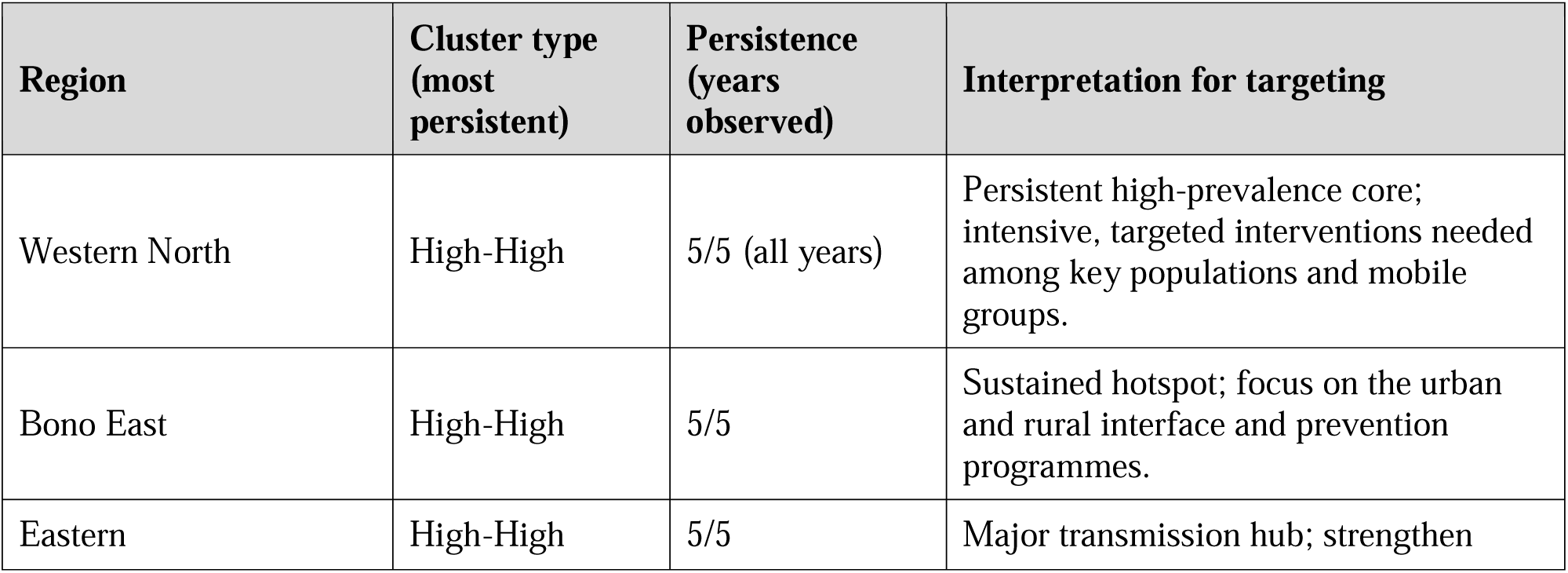
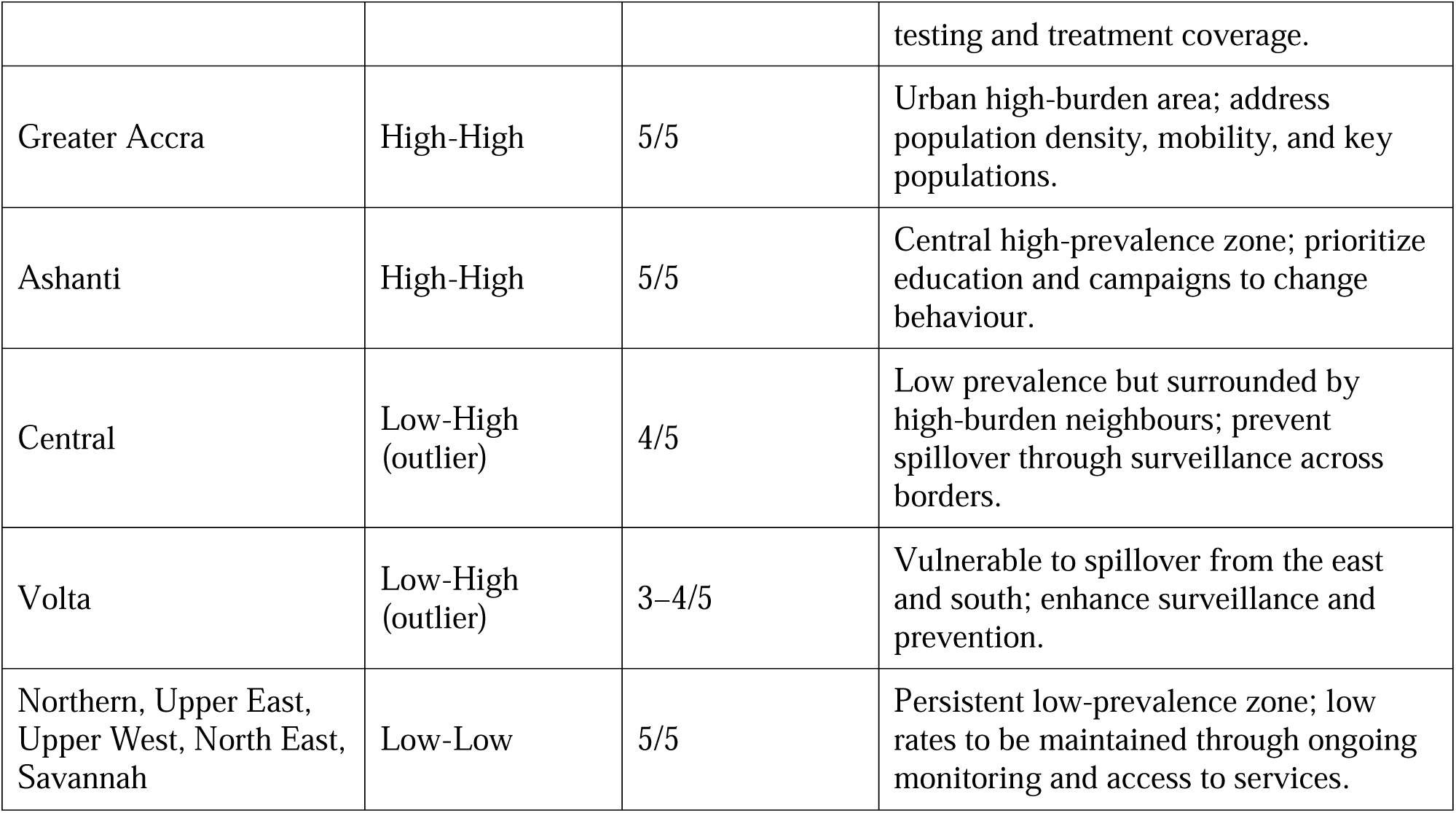
Summary of persistent spatial clustering patterns (local Moran’s I analysis, 2020–2024)

Regions bordering hotspot areas showed consistent outlier status across the study period, a pattern with direct relevance for designing targeted interventions such as surveillance across borders.

## 4.0 Discussion

This study found a general rise in HIV prevalence across Ghana over the study period, alongside substantial variation between regions of high and low burden [30]. Greater Accra, Eastern, and Ashanti recorded consistently high prevalence, while North East and Savannah recorded comparatively low prevalence, a pattern consistent with earlier studies conducted in Ghana [34,35]. The Global Moran’s Index showed no significant spatial pattern in 2021, 2022, or 2023, suggesting that factors other than geographic proximity were likely more important in shaping HIV transmission during this period [31,32]. The Getis-Ord Gi* and local Moran’s I analyses, however, identified local patterns that a global index could not detect: hotspot regions (Ahafo, Ashanti, and Western North) and coldspot regions (Northern, North East, Upper East, Upper West, and Savannah) remained largely consistent across the study period, pointing to stable and geographically coherent clustering over time. The distinction between global and local spatial statistics is well recognized in the spatial epidemiology literature [24,29,32] and is among the reasons this study combined Global Moran’s I with local LISA statistics, an approach suited to disease mapping below the national scale [24].

Several factors may explain the rise in prevalence within high-burden areas, including growing population density in urban centres, greater mobility, and the concentration of key populations such as female sex workers and men who have sex with men [31]. Although the intensity of these hotspots varied somewhat over time, their persistence in southern Ghana [6,7,25] suggests that structural factors, including access to health care, surveillance capacity, urbanization, education, and economic opportunity, continue to shape the country’s HIV landscape.

Lower prevalence in the northern regions may reflect cultural norms that discourage high-risk sexual behaviour, together with lower urbanization and population density [33]. These findings warrant caution, however, because underreporting and limited testing capacity in remote parts of northern Ghana could also produce an apparent, rather than a true, reduction in prevalence. Other studies conducted within Ghana’s national HIV programme have similarly identified gaps in data quality and testing coverage in rural and underserved areas, which may contribute to the north-to-south gradient observed in this study [27,28].

These findings support continued investment in geospatial monitoring to detect shifts in HIV transmission patterns early. Methods such as the Getis-Ord Gi* statistic and both global and local forms of Moran’s I have proved useful for identifying hotspots and can help the Ghana AIDS Commission and Ghana Health Service target interventions and allocate resources more effectively.

### Strengths and Limitations of the Study

This study has several strengths. It drew on routinely collected national HIV surveillance data covering all sixteen regions of Ghana over five years, offering a comprehensive picture of the geographic distribution of prevalence without reliance on survey samples. It applied both global and local spatial autocorrelation methods to identify regions of elevated and reduced burden and to characterize geographic clustering. It also used five years of annual data to assess the consistency and persistence of spatial patterns over time, providing evidence to support surveillance and control strategies targeted by geography.

Several limitations should also be acknowledged. The analysis relied on data aggregated at the regional level, which may obscure variation at the district or community level. Observed spatial patterns may also reflect differences in data quality, testing rates, and reporting practices across regions rather than true differences in transmission, and some apparent patterns may therefore be spurious. Because the study relied solely on cases confirmed by laboratory testing within DHIMS2, it may not fully capture the burden of HIV within the community, particularly where access to testing is limited. Finally, because DHIMS2 does not disaggregate new from existing diagnoses at the regional level, this study examined prevalence rather than incidence; future work using facility-level, line-listed data would allow incidence-based analysis and a more complete picture of transmission dynamics.

### Public Health Implications of the Study

These results support a regional approach to HIV prevention and treatment rather than a uniform national strategy. In high-burden southern areas, intensified prevention, including expanded testing, rapid initiation of antiretroviral therapy, and targeted outreach to key populations, is warranted [27,28]. Peer navigation and mobile testing based in the community have proved effective in other urban centres and should be prioritized [6,25]. Sustained investment in surveillance is needed to prevent escalation in the northern regions, alongside enhanced surveillance across borders in Central and Volta to contain the risk of spillover transmission [2,28].

## 5.0 Conclusions

HIV prevalence in Ghana is rising alongside widening regional inequality, with the more urbanized southern regions recording the greatest burden. Interventions guided by data and tailored to specific regions, including stronger surveillance systems and a more equitable distribution of health resources, are needed to reduce transmission and strengthen Ghana’s HIV control programme.

## Declaration of Generative AI Use

During the preparation of this work, the authors used Claude (Anthropic) to assist with structural revision of the manuscript and with responding to supervisor comments. The authors reviewed and edited the resulting content carefully and take full responsibility for the content of this article.

## Institutional Review Board Statement

Not applicable.

## Informed Consent Statement

Not applicable.

## Data Availability

This study was performed in compliance with relevant laws, regulatory frameworks and guidelines where the research took place.
Ethics committee approval was not required under relevant laws and institutional guidelines. Ethics approval was not required for this study because it involved secondary analysis of routinely collected, aggregated health information obtained from the Ghana District Health Information Management System (DHIMS2). The dataset contained aggregated regional counts and did not include individual-level identifiers or personally identifiable information. The study did not involve direct contact with, recruitment of, or intervention involving human participants. Data were analysed in accordance with applicable institutional and data-governance requirements for the use of routinely collected health information.

https://dhims.chimgh.org/dhims/dhis-web-login/

## Acknowledgements

We acknowledge the Ghana Health Service, the Ministry of Health, and the operational staff of the DHIMS2 programme for the collection and stewardship of these data.

## Author Contributions

O.A.F.I. and F.S.O. conceived and designed the study. O.A.F.I. analyzed the data and wrote the major portions of the manuscript, and F.S.O. wrote the introduction and background. The manuscript was reviewed, edited, and revised for intellectual content by A.H.S., S.D.N., and B.K.Y.A.A. All authors read and approved the final version of the manuscript.

